# Impact of Asian and black donor and recipient ethnicity on the outcomes after deceased donor kidney transplantation in the United Kingdom

**DOI:** 10.1101/2021.05.04.21256445

**Authors:** Abdul R Hakeem, Sonal Asthana, Rachel J Johnson, Chloe Brown, Niaz Ahmad

## Abstract

**Background:** Patients of Asian and black ethnicity face disadvantage on the renal transplant waiting list in the United Kingdom, because of lack of HLA and blood group matched donors from an overwhelmingly white deceased donor pool. This study evaluates outcomes of renal allografts arising from Asian and black donors.

**Methods:** The UK Transplant Registry was analysed for adult deceased donor kidney only transplants performed during January 2001-December 2015.

**Results:** Asian and black ethnicity patients constituted 12.4% and 6.7% of all deceased donor recipients but only 1.6% and 1.2% of all deceased donors, respectively. Across all recipients, and unsurprisingly given the predominantly white recipient pool, HLA matching was superior for grafts from white donors than from Asian and black donors (p<0.0001). Unadjusted survival analysis demonstrated significantly inferior long-term allograft outcomes associated with Asian and black donors, compared to white donors (7-year graft survival 71.9%, 74.0% and 80.5%; log-rank p=0.0007, respectively). On Cox regression analysis, Asian donor (HR 1.37 for Asian donors *vs*. white donors as baseline) and black recipient (HR 1.21 for black recipients *vs*. white recipient as baseline) ethnicities were associated with poorer outcomes than white counterparts, and on ethnicity matching, compared with the white donor–white recipient baseline group and adjusting for other donor and recipient factors, 5-year graft outcomes were significantly poorer for black donor-black [HR 1.92 (1.11-3.32), p=0.02], Asian donor-white recipient [HR 1.56 (1.09-2.24), p=0.016] and white donor-black recipient [HR 1.22 (1.05-1.42), p=0.011] combinations in decreasing order of worse unadjusted 5-year graft survival.

**Conclusions:** Increased deceased donation among ethnic minority communities would benefit the entire recipient pool by increasing the numbers of available organs and may specifically benefit the Asian and black recipients by increasing the numbers of blood group and HLA-compatible grafts for allocation but may not improve allograft outcomes.

## Introduction

United Kingdom (UK) residents of Asian and black ethnicity constitute 14% of the general population (based on 2011 Census estimate), but constitute 20.7% of the total dialysis population, and 32% of the patients on the renal transplant waiting list [1-3]. Poor access to and utilisation of transplant services by ethnic minority population has been well documented in the UK and elsewhere [4,5]. There is a substantial lack of non-white deceased organ donors in the UK donor pool. Whilst organ donation from Asian and black ethnic minorities has significantly increased in the last decade, this increase is offset by the increase in the waiting list patients from these ethnicities. Currently, Asian and black ethnicity contribute 5% of all deceased donation in the UK [3]. Deceased donor kidney allocation within the UK is based on ABO-compatibility and incorporates human leukocyte antigen (HLA) matching between donor and recipient. This puts Asian and black ethnicity recipients at a disadvantage due to the relative scarcity of blood group ‘B’ donors, as well as by challenges in optimal HLA matching with white donors [6-8], although kidney allocation changes made in 2019 sought to minimise disadvantages arising from HLA matching [9]. Significant prevalence of homozygosity of HLA alleles in these populations acts as an additional confounder [10]. In 2019/20 financial year in the UK, 10% of deceased organ donors were blood group B, compared with 19% on the renal transplant waiting list [3]. Despite efforts to improve education about transplant and organ donation among ethnic minorities, awareness remains low [11,12].

Whilst an increase in deceased organ donation from Asian and black ethnicities is desirable to improve access by improving blood group and HLA matching for these recipients, the impact of using organs from minority donors is poorly reported, in part, because of relative scarcity of such transplants. International experience with the use of non-white donors for non-white recipients, has suggested that long-term outcomes are consistently inferior to allografts obtained from white donors [13,14]. There are multiple factors that may contribute to inferior outcome in these settings, in particular higher prevalence of hypertension, diabetes, coronary artery disease and renal disease in these populations [15,16]. The current registry analysis was conducted to compare outcomes of deceased donor allografts derived from Asian, black and white donors in recipients of different ethnicities.

## Materials and Methods

All adult patients who had undergone deceased donor kidney-only transplantation in the UK between January 1, 2001 and December 31, 2015 were eligible for analysis as part of this study (21,206 transplants). For the purposes of this study, ‘Asian’ ethnicity was defined as people of Indian, Pakistani, Bangladeshi or Sri Lankan origin as recorded in the United Kingdom Transplant Registry (UKTR). ‘Black’ ethnicity was defined as people of black, African, Caribbean and black British origin. Patients who received grafts from living donors, paediatric recipients and multiorgan recipients were excluded from the analysis. Also excluded were the transplants where either the donor or recipient ethnicity was not white, Asian or black, or where the recipient gender or HLA mismatch were unknown. Following ethical approval, data were collected from the prospective UK Transplant Registry (UKTR) maintained by NHS Blood and Transplant (NHSBT), on specific donor and recipient variables in addition to graft outcomes.

Donor variables studied were donor ethnicity, age, gender, blood group, and cause of death. Donors were also categorised as extended or standard criteria donors (ECD or SCD). ECDs were those more than 60 years of age, or those aged 50-60 years with at least two of the following risk factors: death due to a cerebrovascular accident, history of hypertension or serum creatinine >1.5 g/dL. Both donors after brain death (DBD) and donors after circulatory death (DCD) were categorised in this way, as there was no evidence of poorer outcomes associated with DCD donors in the UK during this time period [17]. Recipient variables analysed were recipient age, gender, blood group, waiting time to transplant and ethnicity (defined as white, Asian and black ethnicities). Additional data studied included diabetic nephropathy, year of transplant, dialysis status at registration, graft number, cold ischaemia time (CIT) and HLA mismatch (MM) of the transplant (according to the four levels defined for kidney allocation in the UK): Level 1: 000 HLA-A, B, DR MM; Level 2: [0 DR+0/1 B MM]; Level 3: [0 DR+2 B MM] or [1 DR+0/1 B MM]; level 4: [2 B+1DR MM] or [2 DR MM] [18]. In addition, to study the impact of the level of deprivation and ethnicity, recipients were categorised into six different groups based on ‘A Classification of Residential Neighbourhood’ (ACORN) geo-demographic segmentation, which gives us the demographic levels within the UK based on postcodes [19].

### Statistical analysis

Demographic and other factors were analysed for donors as well as for all recipients. White, Asian and black donor characteristics were compared using Chi-squared tests for categorical data and two-tailed t-tests for continuous variables. Data are presented as percentages, or as mean ± standard error, unless otherwise specified.

Graft survival was the primary outcome measure. Graft survival time was death-censored and defined as time from transplant to graft failure. Kaplan– Meier survival curves were used to illustrate differences in graft outcomes. Associated p-values were derived from the univariate log-rank test. Variables were further analysed using Cox proportional hazards regression to determine risk factors for graft failure. The interaction of donor-recipient ethnicity was tested, to assess the effect of different donor-recipient combinations on graft outcome. Results of the Cox regression analysis are presented as estimated hazard ratios (HRs) of groups of individuals compared with that of a baseline group. An HR of greater or less than 1.0 indicates, respectively, a higher or lower risk of failure than in the baseline group. Ninety-five percent confidence intervals (CIs) were calculated for each HR. Log cumulative hazard plots showed no evidence of non-proportionality of hazards.

A 5% level of significance was used, and all analyses were performed using the SAS software package (Version 9.1.3).

## Results

Of the 21,206 transplants from white, Asian or black donor or recipient ethnicity, we excluded 869 (4.1%) transplants that did not have recipient gender or HLA mismatch recorded. This gave 20,337 transplants for final analysis. A further 33 (0.2%) transplants were excluded from the Cox regression analysis due to missing data for recipient waiting time or graft survival time. The analysis cohort of 20,304 transplants from 12,162 donors thus represents 95.7% of all deceased donor kidney only transplants performed in adults in the UK over the study period. Asian (N=195) and black (N=140) donors constituted 1.6% and 1.2%, respectively, of the donor cohort, and the remaining 97.2% were white (N=11,827) donors.

Comparison of white, Asian and black donors (Table 1) showed that black donors were significantly younger (40.5±1.4 years) when compared with the white (47.7±0.1 years) and Asian (45.8±1.2 years) donors (p<0.0001). The Asian and black donors had a significantly different blood group distribution, with higher proportions of blood group ‘B’ (30.8% and 21.4% respectively) and ‘AB’ (7.2% and 3.6% respectively) when compared with white donors (9.1% and 3.3%) (p<0.0001). There were significantly more ECDs among the white donors (32.4%), when compared to Asian (29.2%) and black (20.0%) donors (p=0.0052) and also significantly more DCD donation (29.0% *vs*. 26.7% *vs*. 19.3% respectively; p=0.032). Gender distribution and the incidence of non-traumatic intracranial event as the cause of death were similar in all three groups.

**Table 1:**
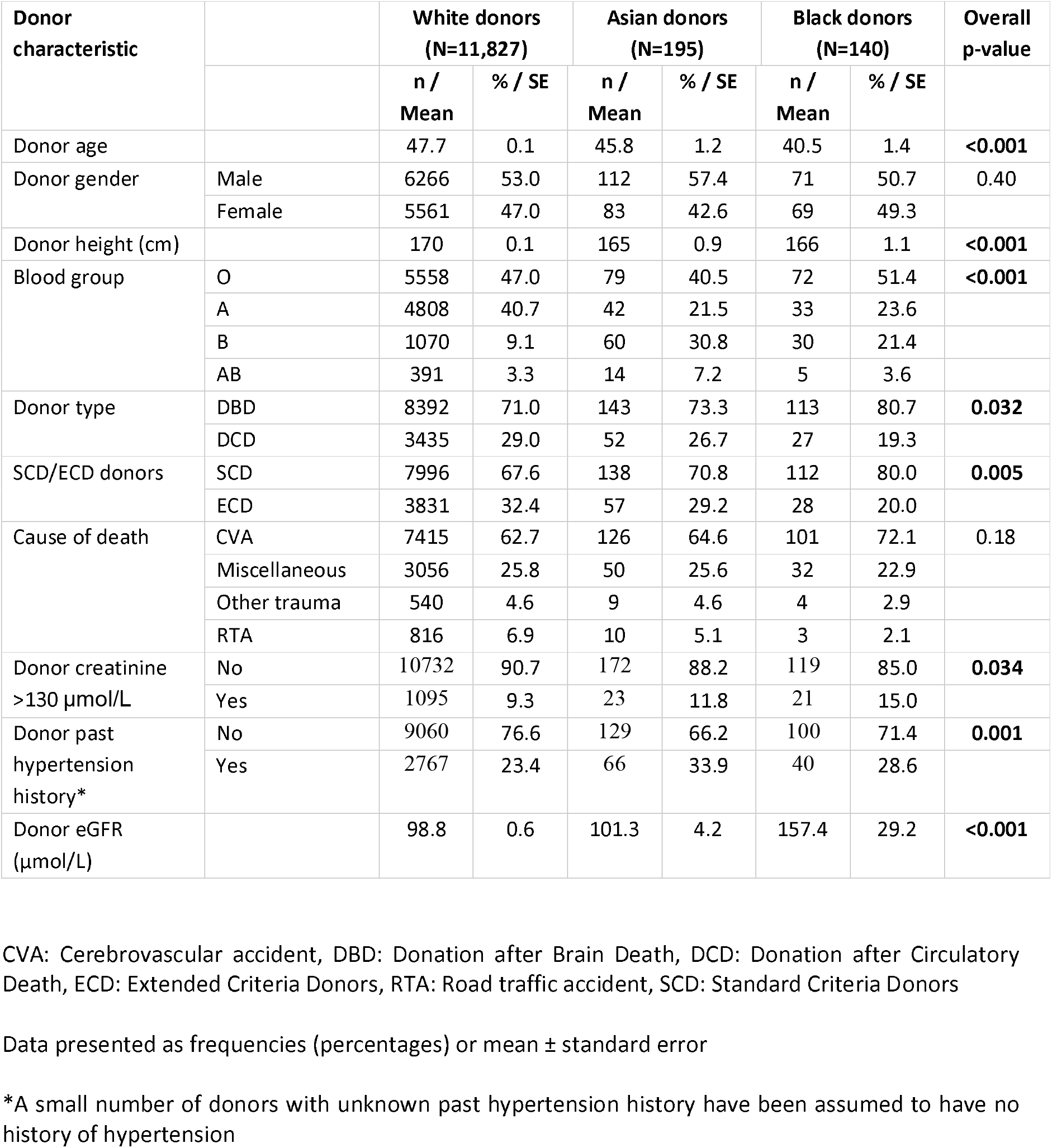
Demographic characteristics of white, Asian and black deceased donors in the UK during 2001-2015 (N=12,162).

| Donor characteristic |  | White donors<br>(N=11,827) |  | Asian donors<br>(N=195) |  | Black donors<br>(N=140) |  | Overall<br>p-value |
| --- | --- | --- | --- | --- | --- | --- | --- | --- |
|  |  | n /<br>Mean | % / SE | n /<br>Mean | % / SE | n /<br>Mean | % / SE |  |
| Donor age |  | 47.7 | 0.1 | 45.8 | 1.2 | 40.5 | 1.4 | **<0.001** |
| Donor gender | Male | 6266 | 53.0 | 112 | 57.4 | 71 | 50.7 | 0.40 |
|  | Female | 5561 | 47.0 | 83 | 42.6 | 69 | 49.3 |  |
| Donor height (cm) |  | 170 | 0.1 | 165 | 0.9 | 166 | 1.1 | **<0.001** |
| Blood group | O | 5558 | 47.0 | 79 | 40.5 | 72 | 51.4 | **<0.001** |
|  | A | 4808 | 40.7 | 42 | 21.5 | 33 | 23.6 |  |
|  | B | 1070 | 9.1 | 60 | 30.8 | 30 | 21.4 |  |
|  | AB | 391 | 3.3 | 14 | 7.2 | 5 | 3.6 |  |
| Donor type | DBD | 8392 | 71.0 | 143 | 73.3 | 113 | 80.7 | **0.032** |
|  | DCD | 3435 | 29.0 | 52 | 26.7 | 27 | 19.3 |  |
| SCD/ECD donors | SCD | 7996 | 67.6 | 138 | 70.8 | 112 | 80.0 | **0.005** |
|  | ECD | 3831 | 32.4 | 57 | 29.2 | 28 | 20.0 |  |
| Cause of death | CVA | 7415 | 62.7 | 126 | 64.6 | 101 | 72.1 | 0.18 |
|  | Miscellaneous | 3056 | 25.8 | 50 | 25.6 | 32 | 22.9 |  |
|  | Other trauma | 540 | 4.6 | 9 | 4.6 | 4 | 2.9 |  |
|  | RTA | 816 | 6.9 | 10 | 5.1 | 3 | 2.1 |  |
| Donor creatinine<br>>130 µmol/L | No | 10732 | 90.7 | 172 | 88.2 | 119 | 85.0 | **0.034** |
|  | Yes | 1095 | 9.3 | 23 | 11.8 | 21 | 15.0 |  |
| Donor past<br>hypertension<br>history* | No | 9060 | 76.6 | 129 | 66.2 | 100 | 71.4 | **0.001** |
|  | Yes | 2767 | 23.4 | 66 | 33.9 | 40 | 28.6 |  |
| Donor eGFR<br>(µmol/L) |  | 98.8 | 0.6 | 101.3 | 4.2 | 157.4 | 29.2 | **<0.001** |

CVA: Cerebrovascular accident, DBD: Donation after Brain Death, DCD: Donation after Circulatory Death, ECD: Extended Criteria Donors, RTA: Road traffic accident, SCD: Standard Criteria Donors
Data presented as frequencies (percentages) or mean ± standard error
\*A small number of donors with unknown past hypertension history have been assumed to have no history of hypertension

Recipient demographics for the 20,337 transplants are presented according to ethnicity of the donor (Table 2). The recipients of kidneys from black donors (47.0±0.9 years) were younger when compared to white (49.7±0.1 years) or Asian (49.1±0.7 years) donors (p=0.010). There were no differences in the proportion of patients with diabetes as the primary diagnosis between the three cohorts. The median waiting time was significantly longer for the recipients who received black (2.8 years) and Asian (2.7 years) donor kidneys, when compared to white (2.2 years) donor kidneys (p<0.0001). Unsurprisingly, kidneys from Asian and black donors were more likely to be transplanted in blood group ‘B’ and ‘AB’ recipients and non-white recipients. The recipients of Asian and black donor kidneys were less likely to be re-graft patients and were less well matched than recipients of white donor organs (Table 2).

**Table 2:** Demographic characteristics of recipients of kidneys from white, Asian and black donors in the UK during 2001-2015 (N=20,337).

| Recipient demographic |  | All recipients of transplants from White donors (N=19,803) |  | All recipients of transplants from Asian donors (N=317) |  | All recipients of transplants from Black donors (N=217) |  | Overall p-value |
| --- | --- | --- | --- | --- | --- | --- | --- | --- |
|  |  | n / Mean | % / SE | n / Mean | % / SE | n / Mean | % / SE |  |
| Recipient age |  | 49.7 | 0.1 | 49.1 | 0.7 | 47.0 | 0.9 | **0.010** |
| Recipient gender | Male | 12412 | 62.7 | 198 | 62.5 | 138 | 63.6 | 0.96 |
|  | Female | 7391 | 37.3 | 119 | 37.5 | 79 | 36.4 |  |
| Blood group | O | 8526 | 43.1 | 112 | 35.3 | 105 | 48.4 | **<0.001** |
|  | A | 7941 | 40.1 | 70 | 22.1 | 47 | 21.7 |  |
|  | B | 2362 | 11.9 | 110 | 34.7 | 53 | 24.4 |  |
|  | AB | 974 | 4.9 | 25 | 7.9 | 12 | 5.5 |  |
| Ethnicity | White | 16191 | 81.8 | 164 | 51.7 | 101 | 46.5 | **<0.001** |
|  | Asian | 2356 | 11.9 | 116 | 36.6 | 49 | 22.6 |  |
|  | Black | 1256 | 6.3 | 37 | 11.7 | 67 | 30.9 |  |
| Diabetes as primary renal disease | No | 18274 | 92.3 | 292 | 92.1 | 200 | 92.2 | 0.99 |
|  | Yes | 1529 | 7.7 | 25 | 7.9 | 17 | 7.8 |  |
| HLA mismatch level | 1 | 2938 | 14.8 | 32 | 10.1 | 16 | 7.4 | **<0.001** |
|  | 2 | 7124 | 36.0 | 94 | 29.7 | 77 | 35.5 |  |
|  | 3 | 8311 | 42.0 | 154 | 48.6 | 105 | 48.4 |  |
|  | 4 | 1430 | 7.2 | 37 | 11.7 | 19 | 8.8 |  |
| Graft | First | 16822 | 85.0 | 283 | 89.3 | 191 | 88.0 | **0.047** |
|  | Regraft | 2981 | 15.0 | 34 | 10.7 | 26 | 12.0 |  |
| Median waiting time (years) and IQ range |  | 2.2 | (0.9 - 3.9) | 2.7 | (1.0 - 4.6) | 2.8 | (1.4 - 4.3) | **<0.001** |
| Median cold ischaemia time (hours) and IQ range |  | 16.0 | (13.0 - 19.7) | 15.3 | (12.6 - 19.0) | 15.8 | (12.2 - 21.0) | 0.14 |
| Recipient Dialysis status at registration | Haemodialysis | 7742 | 39.1 | 120 | 37.9 | 110 | 50.7 | **0.03** |
|  | Peritoneal dialysis | 3856 | 19.5 | 60 | 18.9 | 33 | 15.2 |  |
|  | Not on dialysis | 5066 | 25.6 | 96 | 30.3 | 52 | 24.0 |  |
|  | Unknown | 3139 | 15.9 | 41 | 12.9 | 22 | 10.1 |  |
| Recipient ACORN category | Affluent Achievers | 3658 | 18.5 | 46 | 14.5 | 33 | 15.2 | **<0.001** |
|  | Rising Prosperity | 1195 | 6.0 | 33 | 10.4 | 29 | 13.4 |  |
|  | Comfortable | 5202 | 26.3 | 74 | 23.3 | 48 | 22.1 |  |
|  | Communities |  |  |  |  |  |  |  |
|  | Financially Stretched | 5029 | 25.4 | 63 | 19.9 | 43 | 19.8 |  |
|  | Urban Adversity | 3793 | 19.2 | 85 | 26.8 | 56 | 25.8 |  |
|  | Not Reported | 926 | 4.7 | 16 | 5.1 | 8 | 3.7 |  |

HLA: Human Leukocyte Antigen
Data presented as percentages or mean ± standard error unless stated otherwise

Overall, HLA mismatch levels were superior for grafts from white donors than from Asian and black donors (p<0.0001) [Table 2]. 15% of all white donor kidneys were transplanted with 000 HLA-A, B, DR mismatch, compared to only 10% and 7% of Asian and black donor kidneys, respectively. Better HLA matches were achieved when the donor-recipient pair were of the same ethnicity for all three groups, with 000 HLA-A, B, DR mismatch of 17%, 14% and 13% for white, Asian and black ethnicities, respectively [Figure 1]. The mismatch level was poorest (level 4) when white recipients received kidneys from Asian (15%) and black (14%) donors. For each recipient ethnic group, HLA match differed significantly according to donor ethnicity (p<0.01).

**Figure 1:**
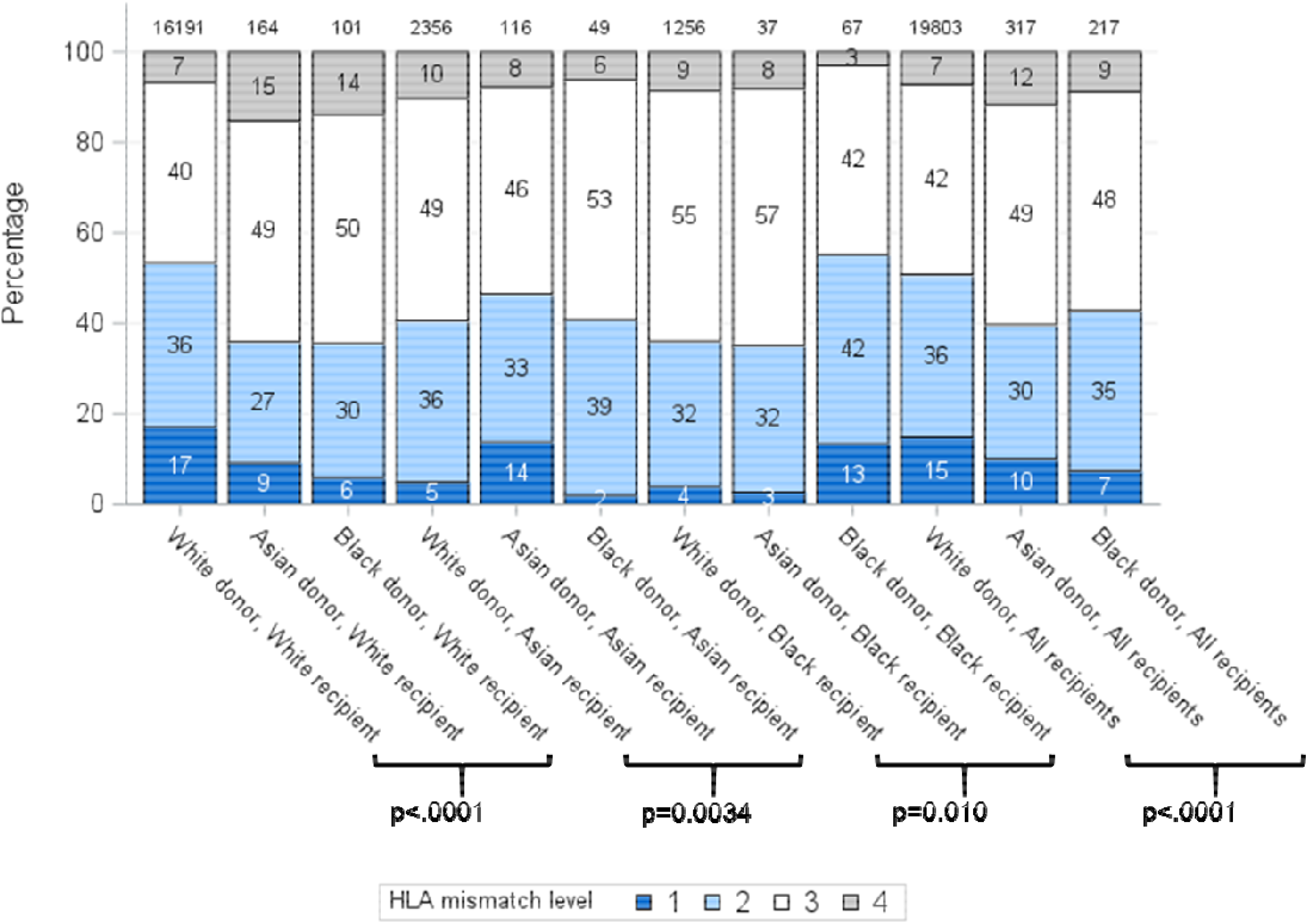
HLA mismatch levels for white, Asian and black donor and recipient combinations. Level 1: 000 HLA-A, B, DR MM Level 2: [0 DR+0/1 B MM] Level 3: [0 DR+2 B MM] or [1 DR+0/1 B MM] Level 4: [2 B+1DR MM] or [2 DR MM] [20]

Unadjusted survival analysis demonstrated significantly inferior long-term allograft outcome for Asian and black donor kidney transplants compared to white donors (7-year graft survival 71.9%, 74.0% and 80.5%; log-rank p=0.0007, respectively) [Figure 2]. Interestingly, further analysis revealed that survival outcomes were worse for black recipients who received grafts from black donors, as compared to kidneys from white donor or Asian donor (7-year graft survival black donor-black recipient 69.2%, compared to white donor-black recipient 74.0%, and Asian donor-black recipient 77.3%, respectively) [Figure 3]. The graft survival rates across donor-recipient ethnicity combinations differed significantly at 3-year (p=0.002), 5-year and 7-year follow-up (p<0.0001), with black donor-black recipient grafts faring worse than all other donor-recipient combinations [Figure 3].

**Figure 2:**
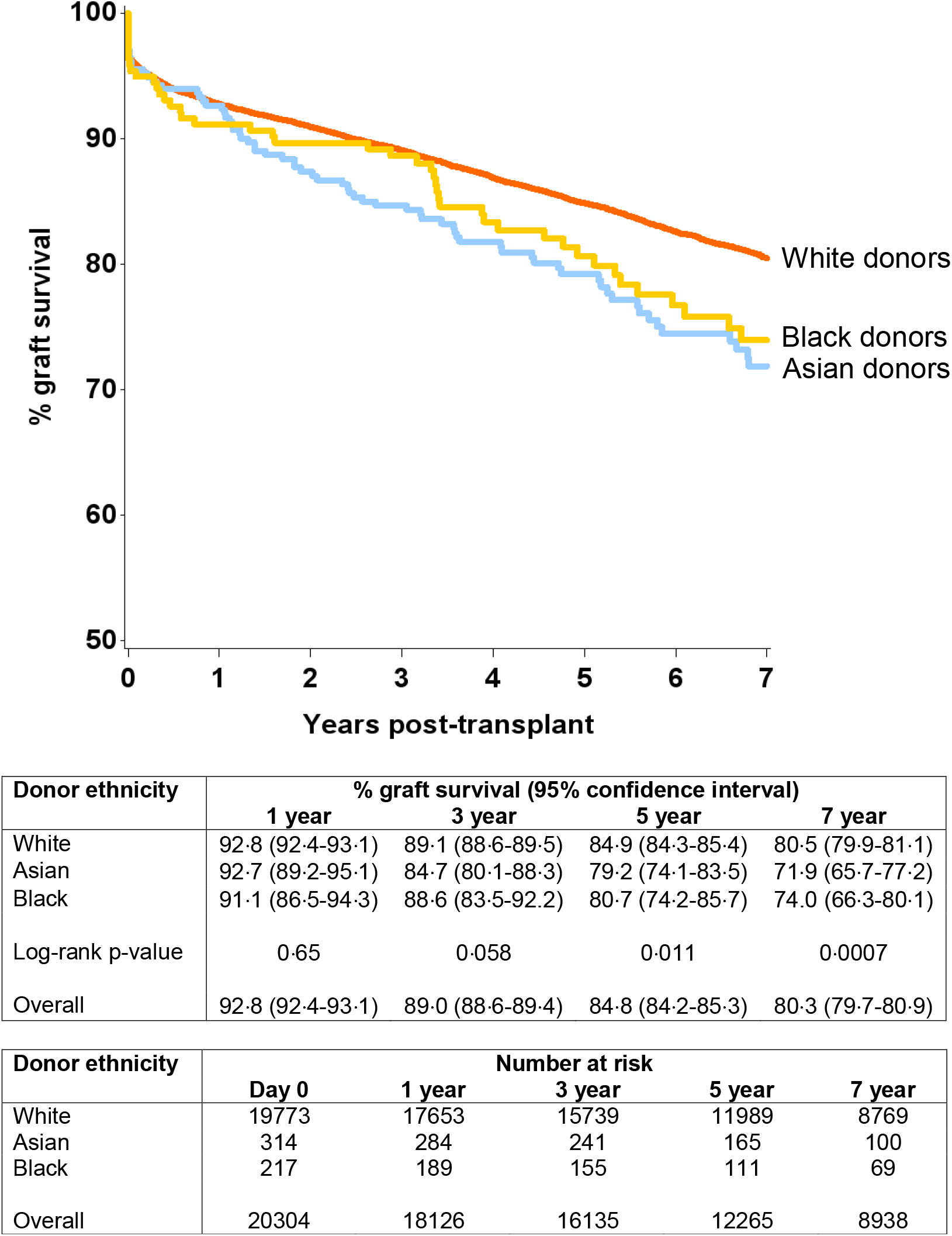
Unadjusted graft survival by donor ethnicity (seven years)

**Figure 3:**
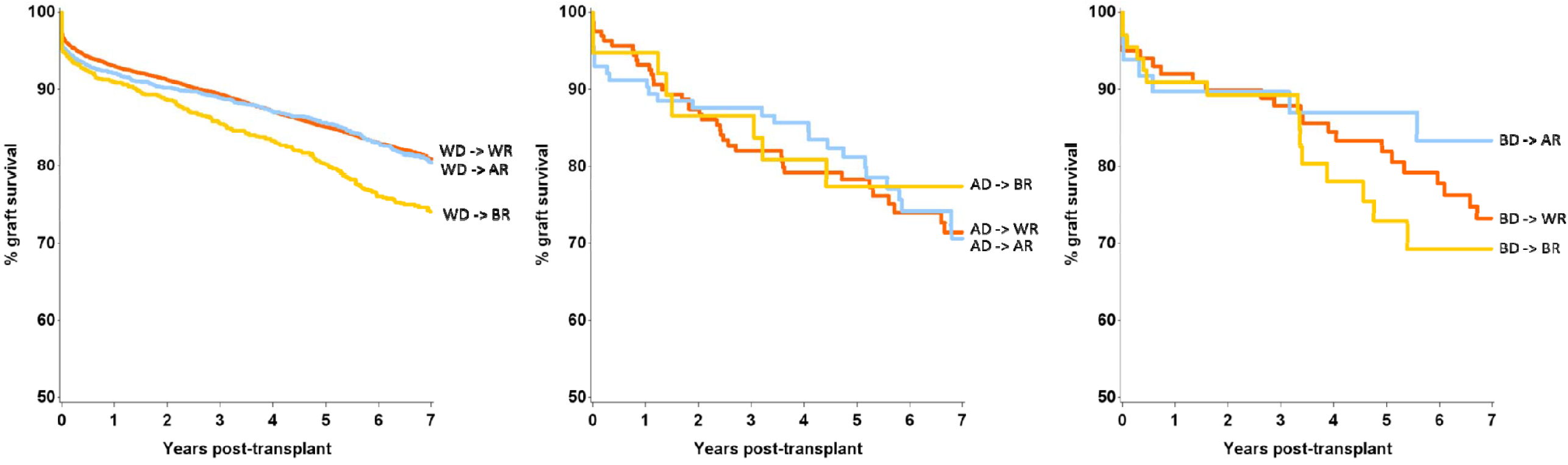
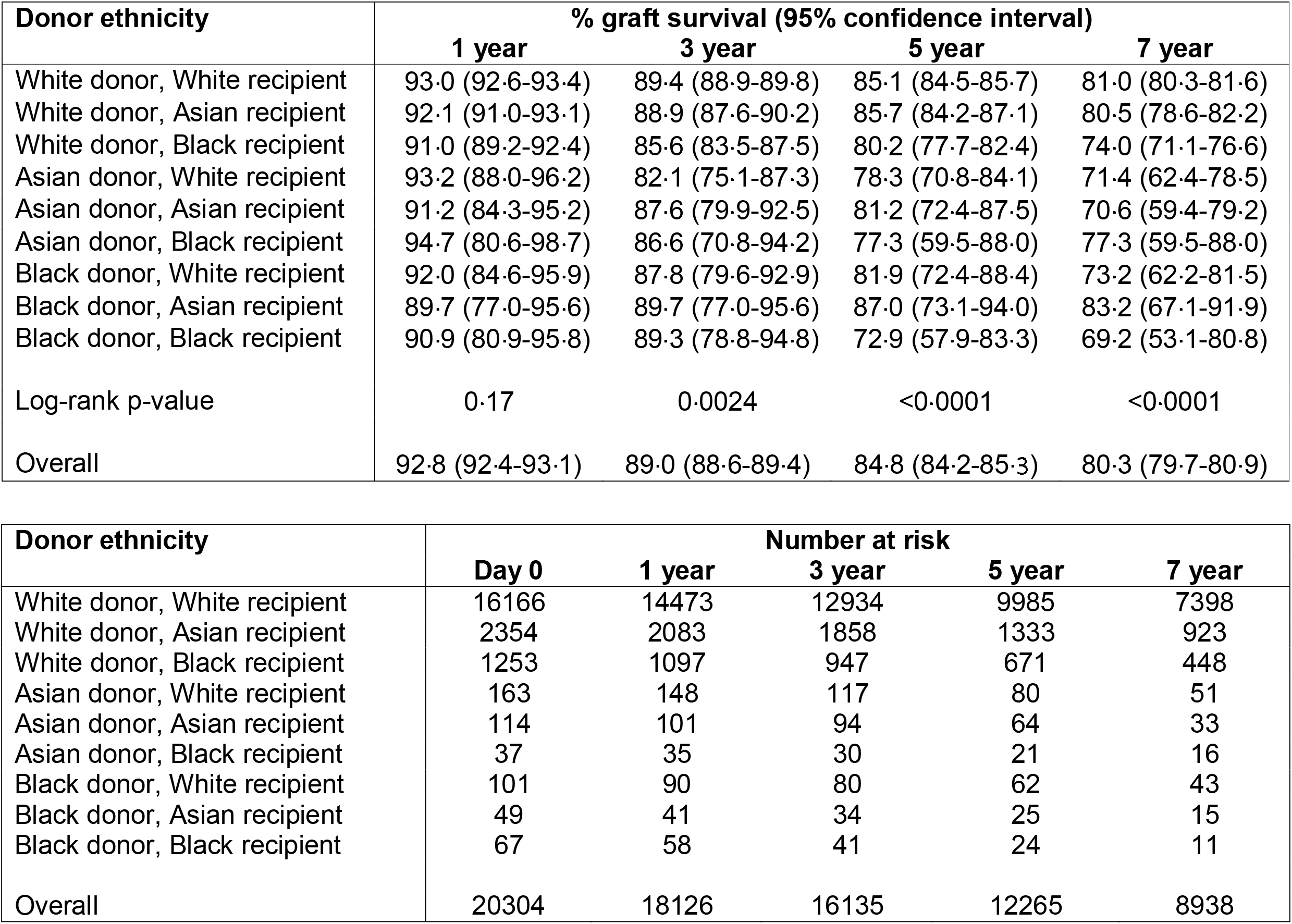
Effect of donor-recipient ethnicity combinations upon graft survival.

Multivariable analysis was performed using the Cox proportional hazards regression model (Table 3). Donor factors associated with 5-year graft failure were age (HR 1.02 for each additional year), male gender (HR 1.0 vs female 0.86), donor height (HR 0.99 for every cm increase in height), donor ethnicity (HR 1.37 for Asian donors *vs*. white donors as baseline), type of donor (HR 1.11 for DCD donors), donor creatinine (HR 1.26 for Cr >130 µmol/L), donor history of hypertension (HR 1.16) and CVA as cause of death (HR 1.12). Recipient factors found to significantly predict graft failure were age (HR 0.78 for each additional year over 60 years), ethnicity (HR 1.21 for black recipients *vs*. white recipient as baseline), dialysis status at transplant (HR 0.88 for peritoneal dialysis and 0.73 for not being on dialysis vs. on haemodialysis as baseline) and waiting time (HR 1.03 for each year of waiting time). Repeat graft (HR 1.37), HLA mismatch (increasing HR for higher levels of HLA mismatch), transplant year (HR 0.96) and cold ischaemia time (HR 1.01 for each minute increase) were also statistically significant (Table 3). The recipient ACORN categories including comfortable communities (HR 1.10), rising prosperity (1.11), financially stretched (HR 1.30) and urban adversity (1.32) showed increasing HR for graft loss, compared with affluent achievers as baseline.

**Table 3:** Cox regression analysis of the donor and recipient factors influencing 5-year graft survival (N=20,304).

| Factor | Level | N | HR | (95% CI) | P-value | Overall p-value for factor |
| --- | --- | --- | --- | --- | --- | --- |
| DONOR |  |  |  |  |  |  |
| Donor age (years) |  | 20304 | 1.02 | (1.02 - 1.02) | <.0001 | **<0.001** |
| Donor gender | Male | 10783 | 1 |  |  | **<0.001** |
|  | Female | 9521 | 0.86 | (0.78 - 0.94) | 0.0008 |  |
| Donor height (cm) |  | 20304 | 0.99 | (0.98 - 0.99) | <.0001 | **<0.001** |
| Donor ethnicity | White | 19772 | 1 |  |  | **0.04** |
|  | Asian | 314 | 1.37 | (1.04 - 1.79) | 0.023 |  |
|  | Black | 218 | 1.28 | (0.91 - 1.80) | 0.16 |  |
| Donor type | DBD | 14338 | 1 |  |  | **0.04** |
|  | DCD | 5966 | 1.11 | (1.01 - 1.23) | 0.039 |  |
| Donor creatinine >130 µmol/L | No | 18380 | 1 |  |  | **0.001** |
|  | Yes | 1924 | 1.26 | (1.10 - 1.44) | 0.0008 |  |
| Donor past hypertension history | No | 15423 | 1 |  |  | **<0.001** |
|  | Yes | 4881 | 1.16 | (1.06 - 1.27) | 0.0008 |  |
| Donor CVA as cause of death | No | 7378 | 1 |  |  | **0.01** |
|  | Yes | 12926 | 1.12 | (1.02 - 1.22) | 0.013 |  |
| Donor eGFR (10 µmol/L) |  | 20304 | 0.99 | (0.98 - 1.00) | 0.1 | **0.07** |
| RECIPIENT |  |  |  |  |  |  |
| Recipient age (years) | 18-39 | 4837 | 1 |  |  | **<0.001** |
|  | 40-59 | 10141 | 0.72 | (0.65 - 0.79) | <.0001 |  |
|  | 60+ | 5326 | 0.78 | (0.70 - 0.88) | <.0001 |  |
| Recipient ethnicity | White | 16430 | 1 |  |  | **0.03** |
|  | Asian | 2517 | 0.98 | (0.87 - 1.11) | 0.76 |  |
|  | Black | 1357 | 1.21 | (1.05 - 1.41) | 0.0097 |  |
| Recipient ACORN category | Affluent Achievers | 3726 | 1 |  |  | **<0.001** |
|  | Rising Prosperity | 1264 | 1.11 | (0.93 - 1.34) | 0.25 |  |
|  | Comfortable Communities | 5298 | 1.10 | (0.97 - 1.24) | 0.15 |  |
|  | Financially Stretched | 5102 | 1.30 | (1.15 - 1.47) | <.0001 |  |
|  | Urban Adversity | 3949 | 1.32 | (1.16 - 1.50) | <.0001 |  |
|  | Other/Not Reported | 965 | 1.03 | (0.84 - 1.26) | 0.75 |  |
| Recipient Dialysis status at registration | Haemodialysis | 7959 | 1 |  |  | **<0.001** |
|  | Peritoneal dialysis | 3943 | 0.88 | (0.80 - 0.98) | 0.019 |  |
|  | Not on dialysis | 5206 | 0.73 | (0.65 - 0.81) | <.0001 |  |
|  | Unknown | 3196 | 0.89 | (0.79 - 1.01) | 0.067 |  |
| Waiting time (years) |  | 20304 | 1.03 | (1.01 - 1.05) | 0.0008 | **0.001** |
| Graft number | First transplant | 17269 | 1 |  |  | **<0.001** |
|  | Re-transplant | 3035 | 1.37 | (1.24 - 1.52) | <.0001 |  |
| HLA mismatch level | 1 | 2995 | 1 |  |  | **0.01** |
|  | 2 | 7320 | 1.11 | (0.98 - 1.25) | 0.097 |  |
|  | 3 | 8531 | 1.20 | (1.05 - 1.36) | 0.0055 |  |
|  | 4 | 1458 | 1.31 | (1.10 - 1.56) | 0.0029 |  |
| Transplant year |  | 20304 | 0.96 | (0.95 - 0.98) | <.0001 | **<0.001** |
| Cold ischaemia time (hrs) |  | 20304 | 1.01 | (1.00 - 1.02) | 0.0012 | **0.001** |

ACORN: Association of Community Organisations for Reform Now, CI: Confidence interval, CVA: Cerebrovascular accident, DBD: Donation after brain death, DCD: Donation after circulatory death, eGFR: estimated glomerular filtration rate, HLA: Human Leukocyte Antigen, HR: Hazard ratio

Further modelling investigated the donor-recipient ethnicity interaction adjusted for all other significant factors (excluding main effects for donor and recipient ethnicity) (Table 4). This showed significantly poorer outcomes compared with the baseline group (white donor-white recipient) for a white donor-black recipient combination [HR 1.22 (1.05-1.42), p=0.011], for Asian donor-white recipient combination [HR 1.56 (1.09-2.24), p=0.016] and for black donor-black recipient combination [HR 1.92 (1.11-3.32), p=0.02]. On comparison of graft survival for donor-recipient pairs of the same ethnicities, the white donor-white recipient pair did significantly better than the Asian-Asian and black-black donor and recipient pairs at 7-year follow-up (81.0% vs. 70.6% and 69.2%, p=0.017). This disparity was not significant over the first three years post-transplant, after which time the survival curves started to diverge until the end of the study period (seven years). [Figure 4].

**Table 4:** Cox regression analysis of donor-recipient ethnicity influencing 5-year graft survival, adjusted for all factors shown in Table 3 except donor and recipient ethnicity (N=20,304).

| Donor-recipient ethnicity | N | Number of events | HR | (95% CI) | P-value |
| --- | --- | --- | --- | --- | --- |
| White donor, White recipient | 16166 | 2129 | 1 |  |  |
| White donor, Asian recipient | 2354 | 299 | 0.99 | (0.874 - 1.12) | 0.86 |
| White donor, Black recipient | 1253 | 200 | 1.22 | (1.05 - 1.42) | **0.011** |
| Asian donor, White recipient | 163 | 30 | 1.56 | (1.09 - 2.24) | **0.016** |
| Asian donor, Asian recipient | 114 | 18 | 1.28 | (0.80 - 2.04) | 0.30 |
| Asian donor, Black recipient | 37 | 7 | 1.19 | (0.56 - 2.50) | 0.65 |
| Black donor, White recipient | 101 | 15 | 1.17 | (0.70 - 1.95) | 0.55 |
| Black donor, Asian recipient | 49 | 6 | 1.07 | (0.48 - 2.39) | 0.87 |
| Black donor, Black recipient | 67 | 13 | 1.92 | (1.11 - 3.32) | **0.02** |

**Figure 4:**
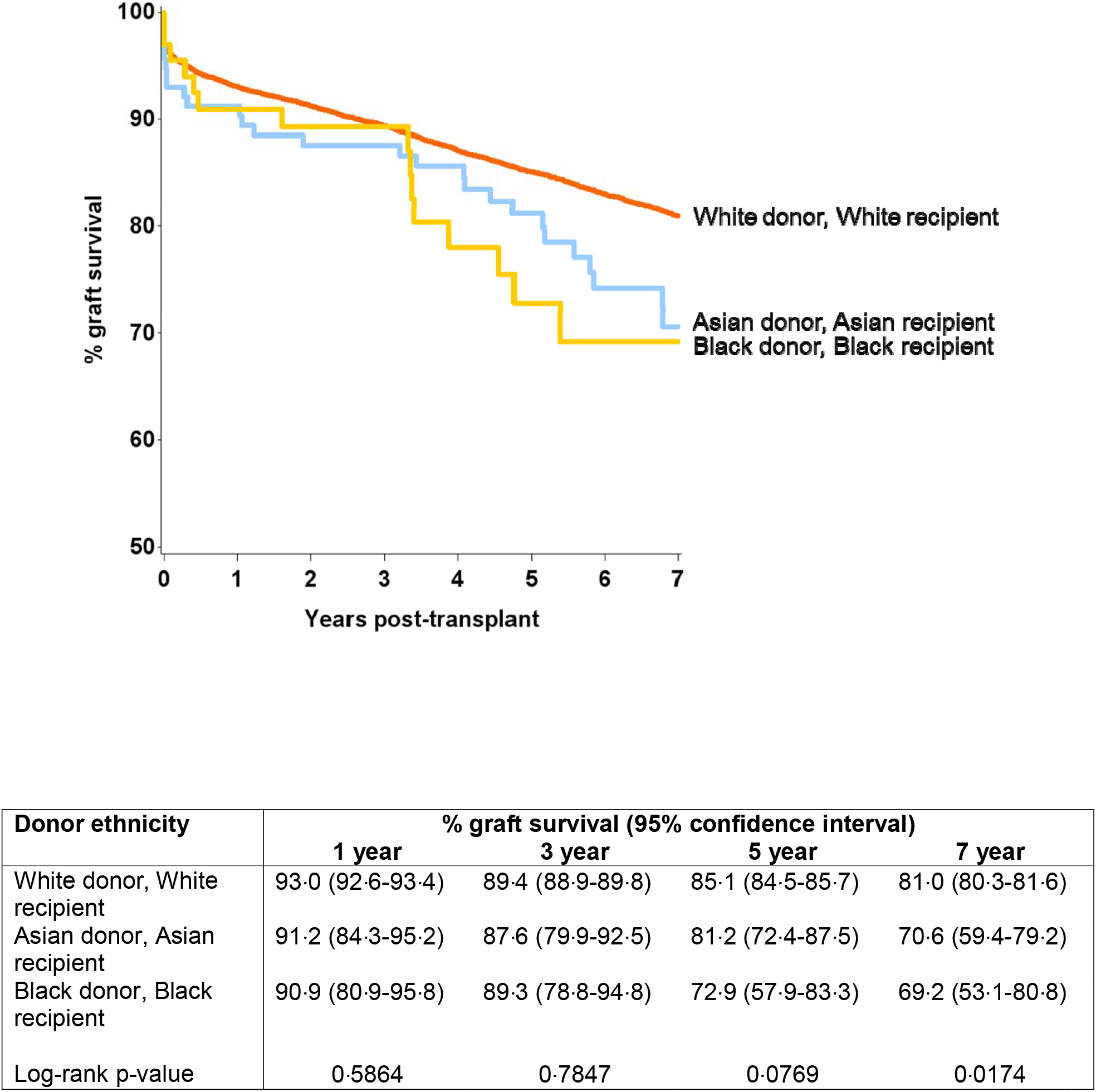
Graft survival from donors of same ethnicity (black-black and asian-asian donor and recipient combination were associated with inferior graft survival at seven year)

## Discussion

This registry analysis, conducted to examine the impact of donor-recipient ethnicity on the outcomes of deceased donor adult renal transplantation in the UK, demonstrated significantly worse graft outcomes associated with Asian donors and black recipients. When compared with white donor-white recipient combination, significantly poorer graft outcomes were observed for black donor-black recipient, Asian donor-white recipient and white donor-black recipient pairs, in decreasing order of worse 5-year graft survival. Asian and black origin patients constituted 12.4% and 6.7% of all deceased donor recipients over the study period; however only 1.6% of donors were of Asian origin and only 1.2% of donors were of black origin. Organ donation rates from Asian and black ethnicity populations have increased in recent years following sustained campaigns, yet significant disparity persists due to the increasing number of patients on the transplant waiting list from these ethnicities [3]. Black donors were significantly younger and more likely to be DBD donors and of standard criteria. The Asian and black donors had higher proportions of blood group ‘B’ and ‘AB’ individuals as compared to white deceased donors in the study population.

The levels of HLA mismatch for organs from Asian and black donors were significantly higher for the entire recipient pool compared to mismatch for transplants from white donors, but Asian and black recipients had more favourable HLA mismatch for organs from those ethnicities, compared to organs from white donors. Asian donor ethnicity and black recipient ethnicity were predictive of graft loss on multivariable analysis, after accounting for all identified significant factors. Further analysis suggested that black recipients of black donor organs had the poorest graft survival of all combinations (5-year and 7-year graft survival black donor-black recipient 72.9% and 69.2%, in comparison to all other pairs where the graft survival ranged from 77.3 to 87.0% and 70.6 to 83.2%, respectively) [Figure 3].

Despite efforts to improve education about transplant and organ donation among ethnic minority groups, awareness and donation rates remain low, when compared to the white population [7,8,20]. Targeted community interventions have not improved deceased donation rates [12,21,22]. Ethnic minority patients face significant disadvantages in access to the renal transplant waitlist in the UK and may wait twice as long as white recipients for a deceased donor renal transplant [23,24]. Barriers to transplantation include socioeconomic factors, lack of blood group ‘B’ donors and difficulties in achieving HLA matched organs from the predominantly white donor pool [25]. Increased deceased donation among ethnic minority communities would benefit the entire recipient pool by increasing the numbers of available organs and may specifically benefit the Asian and black recipients by increasing the numbers of blood group and HLA-compatible grafts for allocation. Indeed, descriptive comparison of white and Asian donors revealed a threefold higher proportion of B blood group donors among Asian donors; organs from Asian and black donors also had a significantly better HLA mismatch among recipients of the same ethnic background.

We included only donors of white, Asian and black origin, excluding deceased donor renal grafts derived from donors of other ethnicities during the study period. These ethnicities represented 95% of all ethnic minorities on the transplant waiting list in the United Kingdom. Given the significant difference in renal risk factors between disparate populations, donor outcomes are also likely to differ significantly, particularly between Chinese and mixed populations - these transplants were excluded to remove the confounding effect of these heterogeneous groups on outcome analysis [26]. Socioeconomic status is well known to affect the outcome of patients of many different diseases, including transplant patients. Our study shows that patients in the less affluent ACORN categories do have higher graft loss compared to the affluent achievers, which is an indirect assessment of access to transplant services, compliance to immunosuppression medications and regular consultations, which all could impact on long-term graft outcomes. These differences are important public health concerns and demand further study and focused interventions in these high-risk groups as well as awareness among the transplant healthcare professionals taking care of these patients [27-29].

Kidney grafts from Asian and black donors were associated with significantly worse survival than those from white donors. Further analysis revealed that the white recipients fared better with grafts from white donors, when compared to grafts from Asian donors. Conversely, the Asian recipients had poorer outcomes from grafts of their own ethnicity, when compared to white or black donors (not statistically significant). Overall, the black recipients had the worst graft outcomes, with poorest outcomes for transplants from black donors, when compared to white or Asian donors. While the rates of early graft failure were comparable for the three ethnicity matched groups initially, the difference in outcomes becomes evident and persists from the third year onwards.

Poor outcomes for Asian and black donor-recipient combinations are likely related to a combination of donor and recipient factors. First and the foremost factor is the longer time on dialysis and longer wait for transplant. The inequity in access to transplantation in the ethnic minorities is well documented, with Access to Transplantation and Transplant Outcome Measures (ATTOM) study showing reduced access to preemptive listing for Asian and black patients and higher likelihood of being listed after starting dialysis [30,31] Significantly higher prevalence rates of diabetes, hypertension, coronary artery disease and death from CVA (which is one of the independent risk factors for graft loss) have been reported in these ethnic minorities [32]. Racial disparities in medical conditions and access to healthcare services may also exist among kidney donors [33,34]. Ethnic minority recipients may have higher cardiac co-morbidity, or infectious complications such as cytomegalovirus (CMV) or BK virus nephropathy, but racial differences in such post-transplant events have not been well studied [34].

Sensitisation levels are usually higher in ethnic minority recipients, and failure may also be related to antibodies to HLA or unrecognised non-HLA antigens [35]. Worse outcomes have also been reported for African American DBD donor-recipient combinations as compared to white donor-African American recipient groups in US registry data [36]. Minor HLA differences could play a key role in affecting long term transplant outcomes in ethnic minorities and there may be need for more comprehensive typing techniques to bring out these differences [37]. The differences in immunosuppression drug metabolism could also affect long-term outcomes, as black and mixed-race patients demonstrate very high rates of CYP3A5 expression, with a significant impact on tacrolimus pharmacokinetics and hence need for higher dosing algorithms [38].

Deceased donors from ethnic minority populations were less likely to be considered as extended criteria (29.23% of Asian and 20.0% of black donors *vs*. 32.39% of white donors), probably due to the younger age of death of this cohort compared to white donors. Black ethnicity increases risk of graft failure in donor-risk models and inferior graft outcomes for organs from black donors have been well documented in US-based registry data. Asian populations, like black populations, have higher rates of diabetes, hypertension and renal disease than comparable white population cohorts in the UK [39]. An increased prevalence of renal diseases and co-morbidities affecting kidney function in ethnic minority populations is likely to confer added donor risk from these groups. This study supports such a hypothesis.

This study included patients who had undergone a renal transplant in the UK before 2005. Organ allocation policy for DBD donors in the UK changed in 2006, with an emphasis on equity of access, in addition to HLA matching [9]. This policy appears to have improved access to renal transplantation among ethnic minorities; however, advantages have been offset by an increase in the number of patients on the transplant waiting list [3]. Organs from DCD donors continued to be allocated according to local policy, until September 2014. In 2019 a fully integrated DBD and DCD kidney allocation scheme was introduced in the UK, simulations of which predict improvements in the equity of access to transplant across ethnic and blood groups [40].

This study has several limitations inherent to a large registry-based retrospective analysis. The long-term data collection has the risk of under-ascertainment of events such as death or graft loss, which cannot be refuted. Ethnicity was self-reported, and this analysis offers no information on graft outcome in mixed-race recipients. Data on ethnic minority donors consisted of 2.8% of the entire study cohort, although it represents all such available data from the UK over more than a decade.

In conclusion, expanding the organ donor pool by increasing donation rates among ethnic minority groups remains a worthy goal and will improve overall access to transplantation and reduce time spent on waiting list, in particular within the ethnic minority communities. When looking at ethnicity matching between donor and recipient and compared with white-white, graft outcomes were worse for white-black, Asian-white and black-black renal transplants. Despite advantages of blood-group compatibility and improved HLA matching, black recipients of black donor grafts appear to have the poorest outcomes, and this difference cannot be explained by donor factors alone. There is need for future work to factor this into UK kidney allocation system, so as to get the best long-term results. An increase in deceased organ donation from ethnic minorities may improve access to transplantation for these groups, but may not improve allograft outcomes.

## Data Availability

Not available

## Acknowledgements

We are thankful to the NHSBT for providing the data used in the manuscript and to all transplant centres for providing data to the UK national transplant registry.

## Abbreviations

ACORN: A Classification of Residential Neighbourhood
CI: Confidence Interval
CIT: Cold Ischaemia Time
CMV: Cytomegalovirus
DBD: Donation after Brain Death
DCD: Donation after Circulatory Death
ECD: Extended Criteria Donors
HLA: Human Leucocyte Antigen
HR: Hazard Ratio
MM: Mismatch
NHS: National Health Service
NHSBT: NHS Blood and Transplant
NS: Not significant
SAS: Statistical Analysis Software
SCD: Standard Criteria Donors
UK: United Kingdom
UKRR: United Kingdom Renal Registry
UKTR: United Kingdom Transplant Registry
US: United States

